# Gazing Down Has benefits Unrelated To Visual Input

**DOI:** 10.1101/2023.12.20.23300291

**Authors:** Yogev Koren, Noy Goldhamer, Lior Shmuelof, Simona Bar-Haim

## Abstract

Recent reports have revealed that downward gazing enhances postural control. The mechanism underlying this phenomenon is currently unknown, yet there are several plausible explanations. In this study, we attempt to provide evidence to support the hypothesis that this effect is primarily derived from altered visual flow caused by gazing down.

To this end, we quantified standing postural sway of 20 healthy participants and 20 people with stroke who were instructed to stand as still as possible under different conditions: while gazing forward and gazing down, with their eyes open and eyes closed.

Both the horizontal gaze angle and the lack of visual input had a negative effect on participants’ ability to attenuate their body sway. Yet, the effect of gaze angle was constant regardless of the presence or absence of visual input. Also, people with stroke swayed more than their healthy counterparts and were more sensitive to the effect of gaze angle, but not to that of visual input.

The results of this study indicate that downward gazing enhances postural control even in the absence of visual input and do not support our main hypothesis. Also, it seems that the effect of downward gazing on postural control is greater in unstable people (people with stroke) than that observed in healthy adults, which might explain less stable individuals’ tendency to gaze down while walking. Furthermore, these results might suggest that downward gazing behavior does not necessarily indicate an attempt to acquire visual information of any kind but instead serves to modulate some other sensory input helpful for postural control.

## Introduction

The ability to control the motion of the center of mass (COM) with respect to the base of support (BOS) is a fundamental prerequisite for most human activities. This is true for both static situations, in which the BOS is stationary, and dynamic situations in which the BOS is constantly changing. This ability resides under the umbrella term ‘postural control’ [1].

It is generally accepted that somatosensory, vestibular and visual information are integrated and used to control posture (e.g., [2]). That is, these sensory modalities provide information about the body’s position and motion that is used to generate corrective responses to gravitational and other internal and external forces acting on the body. Both the vestibular and the visual organs are in the head, meaning that they provide information about the head’s position and motion, which might be different from the position and motion of the COM. For example, when standing in a stochastic 3D visual space, forward body motion with the head oriented forward will generate a visual flow that is consistent with the flow generated by forward head motion (visual expansion). The same forward body motion with the head rotated to one side will generate visual flow consistent with mediolateral head motion (motion parallax). The central nervous system (CNS) is flexible enough to accommodate such changes and can therefore provide a directionally adequate response with different head-on-neck and eye-in-head positions [3-5]. Such an effect indicates that postural responses are not ‘rigid’; rather, the CNS uses an internal body scheme/representation when interpreting vestibular and retinal signals. This body scheme is thought to rely mainly on somatosensory input [6], and in the example above, proprioceptors (muscle spindles) in the neck muscle were shown to have great impact on the subjective interpretation of the retinal input [7].

Nevertheless, visual input is also a central source of information regarding body posture. The accumulation of visual information is guided by gaze direction, which can have a dramatic effect on the sensory afferent signal/s. Different areas in the surroundings contain different visual cues; their density, distance from the observer, and whether they are stationary or moving can greatly impact the percept of motion [8]. Furthermore, even if the same visual cues are visible in different gaze positions, whether their image is projected onto the central visual field or onto its periphery can significantly affect vection [9].

Sensory input from the muscles controlling the head and eyes (proprioceptive input) can also change with different head and eye positions. A good example is the effect of gaze distance. Compared with long gaze distance, short gaze distance was shown to enhance postural control [10], presumably due to the larger retinal displacement of the image at this distance, i.e. a change in the retinal afference. Contrary to this presumption, Kapoula and Le [11] showed that artificially manipulating the convergence angle of the eyes effaced the effect of viewing distance. These results indicate that the retinal afferent is not the (only) source for the effect of gaze distance on postural control. Instead, it was suggested that when the muscles controlling the eyes are stretched or when they contract, they provide information more appropriate to aligning the eye’s coordinate system with that of the body than the information provided in their ‘slack’ condition (at smaller convergence angles). The effect of muscle ‘slack’ on muscle spindle discharge and the sense of body position has been previously demonstrated in other muscles [12,13].

Overall, different head positions may affect the body’s biomechanics. Such changes can indirectly affect the ability to control the COM, by placing muscles in their slack position, thereby reducing the accuracy of the internal body scheme; alternatively, they can affect control of COM directly, by shifting the projection of the COM closer to the limits of the BOS and/or by increasing the moments around key joints (e.g., the ankle joint).

Although during daily life activities humans often change their gaze position, very few investigations were conducted to determine how this behavior affects postural control. Moreover, observations from these investigations were inconsistent and sometimes conflicting, with various mechanisms proposed to underly the observed effects. For example, Buckley et al. [14] reported that, with matched visual input and gaze distance, upward and downward head position increased postural sway in comparison to forward gazing (FG). Consistent with this report, Vuillerme and Rougier [15] found the same for upward head position with eyes closed. In contrast, Aoki et al. [16,17] reported that sway values with eyes closed were not different between the downward and forward head positions.

Aoki et al. [16] reported that, without matched retinal input, sway values decreased when healthy young participants gazed down, but this was true only for uniped stance, and for biped stance no difference was observed. In contrast, the same group [17] reported that for elderly participants, downward gazing (DWG) increased sway values in comparison to forward gazing (FG), with and without matching gaze distance, but for people with stroke (PwS) the inverse was observed.

In Aoki’s reports, head and eye angles were not separated (rather, participants were simply instructed where to look), which is an important point to note, as both upward and downward eye positions, with the head positioned straight forward, were shown to decrease sway values in comparison with a forward eye position [11,18], but lateral eye position was reported to increase these values [19].

Recently, Koren et al. [20] reported that DWG enhanced postural steadiness of standing and walking younger adults. These authors also found a similar effect with older adults and PwS [21], two populations that excessively rely on visual input and are more likely to gaze down while walking [22,23]. Based on previous literature, they further speculated that DWG enhances postural control primarily through its effect on the visual (retinal) input. In this investigation we attempted to provide evidence to support this speculation. To do so, we tested whether the effect of DWG on postural sway with visual input (eyes open) was different from the effect observed without visual input (eyes closed). Specifically, we hypothesized that the effect of DWG on postural sway with eyes open would be greater than the effect with eyes closed (if such an effect is even be observed). In other words, we expected to find a Vision by (gaze) Angle interaction, a prediction that previous reports had not tested directly.

## Methods

### Participants

Twenty healthy adults and 20 PwS participated in this study. Participants were recruited at the Adi-Negev Rehabilitation Center from among the patients (in- and outpatients), staff, and visitors. All participants provided written consent prior to testing. Participants were men and women between 18 and 85 years old, able to stand with their eyes closed for 30s, able provide consent, and able to follow simple instructions. Prospective participants were excluded if they had a history of a major orthopedic condition (such as total knee replacement), or present acute orthopedic symptoms (such as severe pain due to osteoarthritis). Prospective participants with neurological (other than stroke), degenerative, or other conditions that can affect postural control (such as vertigo, severe visual impairment, etc.) were also excluded from participating. Prospective participants with common age-related conditions, such as controlled type 2 diabetes, hypertension, etc., were allowed to participate. The study was approved, beforehand, by the local ethical review board at Sheba Medical Center, Israel (approval number 6218-19-SMC).

### Procedure

Participants were instructed to stand barefoot, as still as possible, in a standardized wide-base stance, i.e., their heels 6cm apart with the feet externally rotated (10°), and their hands loosely hanging to the sides of their bodies [24]. Their postural sway was measured while gazing forward or downward, with their eyes closed or open, for 30s in each trial. This can be visualized as a 2×2 conditions matrix (2 visual conditions and 2 angle conditions). For the forward condition, participants gazed at a target placed at eye level, located 4 meters ahead, and for the downward condition the target was placed on the standing surface, 2 meters ahead. The gaze targets were colored circles, 20cm in diameter, made of laminated paperboard. In the eyes-open condition, participants were instructed to gaze at the designated target constantly throughout the trial. In the eyes closed conditions, participants were instructed to first gaze at the designated target with their eyes open and then to close their eyes while imagining that they are still gazing at the target. This instruction was specifically provided to match the eyes-in-head position between the eyes-open (EO) and the eyes-closed (EC) conditions.

Five repetitions [25] of each condition were performed in a random order using a Latin rectangle. That is, a 5×4 matrix, in which columns are the testing conditions and rows are the repetitions, was automatically generated for each participant. The order of columns in each row was random and different for each participant. Before and after the execution of the Latin rectangle, a single, free-gaze trial was performed with no targets and the participant was instructed to gaze at his/her preferred angle (with eyes open).

Rest between trials was provided as necessary without any restrictions. Participants that reported using corrective visual aids for usual daily activities (myopia but not presbyopia) were tested with their own aids.

### Instruments

To measure sway during the trials, participants stood on a platform equipped with an embedded force sensor array (Zebris FDM-T Treadmill, *Zebris Medical GmbH*, Germany). For consistency, the standardized foot position was marked on the platform. Raw data were acquired using the software provided by the manufacturer (Zebris FDM, version 1.18.40) at a 60 Hz sampling rate. Since the platform is elevated from the laboratory’s floor, a 1.70×0.7×0.3m wooden platform was custom built to create an illusion of continuity of the standing surface (on which the target for the downward gaze was placed), and participants were tested while facing the back end of the treadmill.

To measure head angle throughout the experiment, participants were fitted with a single inertial measuring unit (IMU) on their forehead. The IMU (Xsens DOT, Movella, Netherlands) was placed in a special case attached to an elastic band, designed for this purpose (provided by the manufacturer). Data from the IMU was sampled at 60 Hz and acquired wirelessly using software provided by the manufacturer (Version 2020.0.1).

### Data processing and outcome measures

Raw data from the force platform was exported and processed by a dedicated MATLAB script. First, the center of pressure (COP) time series was low passed using a 2^nd^ order Butterworth filter with a cutoff frequency of 15 Hz. The script, which excludes the first three seconds and the last second of each trial, computes four traditional sway parameters from the individual time series: COP range in the anterior-posterior (AP) and medio-lateral (ML) directions is simply the distance between the extreme values on the Y and X axes, respectively, and given in mm. Sway velocity, given in mm/sec, was calculated as the total excursion divided by time. The fourth parameter was sway area (given in mm^2^). We chose to calculate the area of the smallest convex set containing all visited points, i.e., the convex hull of the 2D data (as described in [26]).

As the main outcome measure, the script computes the short-term diffusion coefficient (Dis) of COP, driven from stabilogram diffusion analysis (SDA) as described by Collins and De-Luca [27]. Briefly, the diffusion coefficient is the rate at which the quadratic Euclidian distance between two COP positions increases as a function of the time interval between them. Thus, for a given Δt, spanning m data intervals and N samples, planar displacement (Δr^2^) is calculated as:

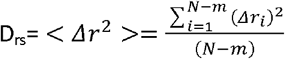

This calculation is repeated for every Δt and the Dis is calculated as the slope of the Δr^2^ by Δt plot over the initial part of the plot (spanning roughly 1 s). In this experiment we calculated three coefficients, all given in mm^2^/sec: the two single dimensions on the X (Dxs) and Y (Dys) axes, and the planar coefficient (Drs).

SDA parameters were shown to be more sensitive than summary statistics (traditional parameters) to postural instability [24]. Nevertheless, many researchers prefer using traditional sway metrics, so we included both. Regardless of the true nature of standing sway (which is debated in the literature), the task in this study required participants to minimize sway as much as possible; therefore, smaller values (of all parameters) are interpreted as a better ability to control the COM.

To determine the vertical head angle in each trial, the data from the force platform and the vertical angle (relative to the gravitational vector) data from the IMU were synchronized using the time stamps from the devices. The head angle was determined as the mean value during the trial.

### Sample size estimate

To estimate the number of participants required to show an effect of DWG on postural sway, we used the data collected from our previous study, which included four participants (one older adult and three PwS) who were tested in a wide-base stance [21]. We used the ‘SIMR’ package [28] in R (Version 4.0.5), in conjunction with the ‘lme4’ package [29]. This package allows users to calculate power for generalized linear mixed models. The power calculations are based on Monte Carlo simulations [28]. We simulated multiple experiments with DWG (to 3 meters ahead) and FG as levels of the fixed effect, at various levels of the random effect (i.e., number of participants). When the predicted term in these simulations was the variable Drs, the observed power reached 92% (CI: 85-96) with 20 participants. Given that we were (mostly) interested in the interaction term (‘Vision’ by ‘Angle’) and not the main effect of the angle, we decided to recruit 20 participants in each group. This value is much greater than that previously estimated [21], and is probably the result of testing participants in a wide-base-stance instead of narrow-base-stance, which is more sensitive to postural instability [30].

### Statistical analysis

In all cases, mixed-effects models were used for the analysis (using SPSS, Version 29, IBM Corp, Armonk, NY). For the main objective, models included participants as the random effect. For fixed effects, we used ‘Group’, ‘Vision’, ‘Angle’, and all possible interactions (full factorial models). Non-significant terms were excluded from the model in a stepwise manner. Significance level was set a-priori at α<0.05, and sequential Bonferroni was used to correct for multiple comparisons where appropriate. For effect size, we used the marginal pseudo R^2^, which quantifies the variance explained by the fixed effects in the model. The distribution of all sway parameters was skewed to the right and therefore adjusted using a logarithmic transformation (natural log). The residuals of all models were evaluated for their distribution.

## Results

One participant did not complete the full experimental protocol due to tiredness. This participant performed only the first free-gaze trial and four trials of each of the other conditions. Data from seven other trials (from three different participants) were excluded from the final analysis due to interruptions (participant either moved or talked, or other disturbance occurred during the trial). Head angle data were missing in five trials (from three participants) due to technical difficulties. Overall, sway data from 868/875 trials, performed by 20 healthy adults and 20 PwS, were used in the final analysis. Participants’ characteristics are presented in Table 1.

|  | Healthy |  | Stroke |  |
| --- | --- | --- | --- | --- |
|  | men | women | men | women |
| N | 10 | 10 | 15 | 5 |
| Height<sub>cm</sub> | 176 [165-185] | 161 [153-170] | 171 [156-184] | 159 [154-167] |
| Weight<sub>kg</sub> | 85 [71-121] | 65 [52-89] | 80 [59-105] | 63 [48-79] |
| BMI<sub>kg/m<sup>2</sup></sub> | 27.3 [23.3-35.3] | 25.3 [19.8-35.8] | 27.3 [20.1-32.6] | 24.9 [20.2-31.2] |
| Age<sub>years</sub> | 49 [26-75] | 44 [25-68] | 61 [30-76] | 71 [64-77] |
| TFO<sub>months</sub> |  |  | 3 [1-26] |  |
| FM-LE |  |  | 23.7 [6-34] |  |
| 10MWT<sub>m/sec</sub> |  |  | 0.8 [0-1.4] |  |

*\*Values represent mean and [range]. TFO is the time (in months) from stroke onset. FM-LE is the motor section of the Fugl-Meyer lower extremity (max score is 34), and the 10MWT is the score (mean walking velocity) in the 10-meter walk test (with 0 meaning that the participant was unable to perform the task).*

### Head angle

First, we wanted to ensure that head angles in the EO and EC conditions were similar, and to estimate whether participants used head-on-neck or eyes-in-head to gaze down. The results of this model revealed that the DWG head angle differed from the FG head angle by roughly 20° (p<0.0001), indicating that DWG was achieved (at least partially) using head-on-neck movement. The mean difference between the EO and EC conditions was <0.5° (DWG=0.57° and FG=0.12°) and was not significant (p>0.39).

### Main results

The results and final statistical models of all sway parameters are presented in Table 2 and in Figures 1-2.

**Table 2.** Final results for all sway outcome measures.

| Parameter | Group | Vision | Angle | G×A | R<sup>2</sup> |
| --- | --- | --- | --- | --- | --- |
| Drs | 1.13 [0.62-1.64]<br>p=1.5×10<sup>-5</sup> | 0.53 [0.44-0.61]<br>p<0.001 | 0.065 [0.021-0.11]<br>p=0.004 |  | 0.32 |
| Dys | 0.99 [0.51-1.47]<br>p=5.1×10<sup>-5</sup> | 0.60 [0.51-0.69]<br>p<0.001 | 0.064 [0.011-0.12]<br>p=0.02 |  | 0.30 |
| Dxs | 1.17 [0.54-1.178]<br>p<0.001 | 0.35 [0.26-0.43]<br>p=2.2×10<sup>-14</sup> | 0.066 [0.011-0.12]<br>p=0.02 | p=0.02 | 0.23 |
| RangeX | 0.53 [0.26-0.80]<br>p<0.001 | 0.094 [0.05-0.14]<br>p=5×10<sup>-5</sup> | 0.001 [-0.030-0.033]<br>p=0.94 | p=0.001 | 0.21 |
| RangeY | 0.41 [0.18-0.63]<br>p<0.001 | 0.17 [0.13-0.26]<br>p=5.1×10<sup>-13</sup> | 0.024 [-0.008-0.057]<br>p=0.14 | p=0.02 | 0.20 |
| Area | 0.93 [0.48-1.39]<br>p=6.9×10<sup>-5</sup> | 0.27 [0.20-0.34]<br>p=5.1×10<sup>-14</sup> | 0.029 [-0.023-0.082]<br>p=0.28 | p=0.003 | 0.25 |
| Velocity | 0.53 [0.28-0.78]<br>p=3×10<sup>-5</sup> | 0.28 [0.24-0.33]<br>p<0.001 | 0.043 [0.023-0.063]<br>p=3.9×10<sup>-5</sup> | p=0.04 | 0.32 |

Values are the mean difference [95% CI] between the two levels of each main (fixed) effect. For the Group by Angle (G×A) interaction, only the p-value is reported. R<sup>2</sup> is the marginal pseudo R<sup>2</sup>, which quantifies the variance explained solely by the fixed effects in the model.

**Figure 1.**
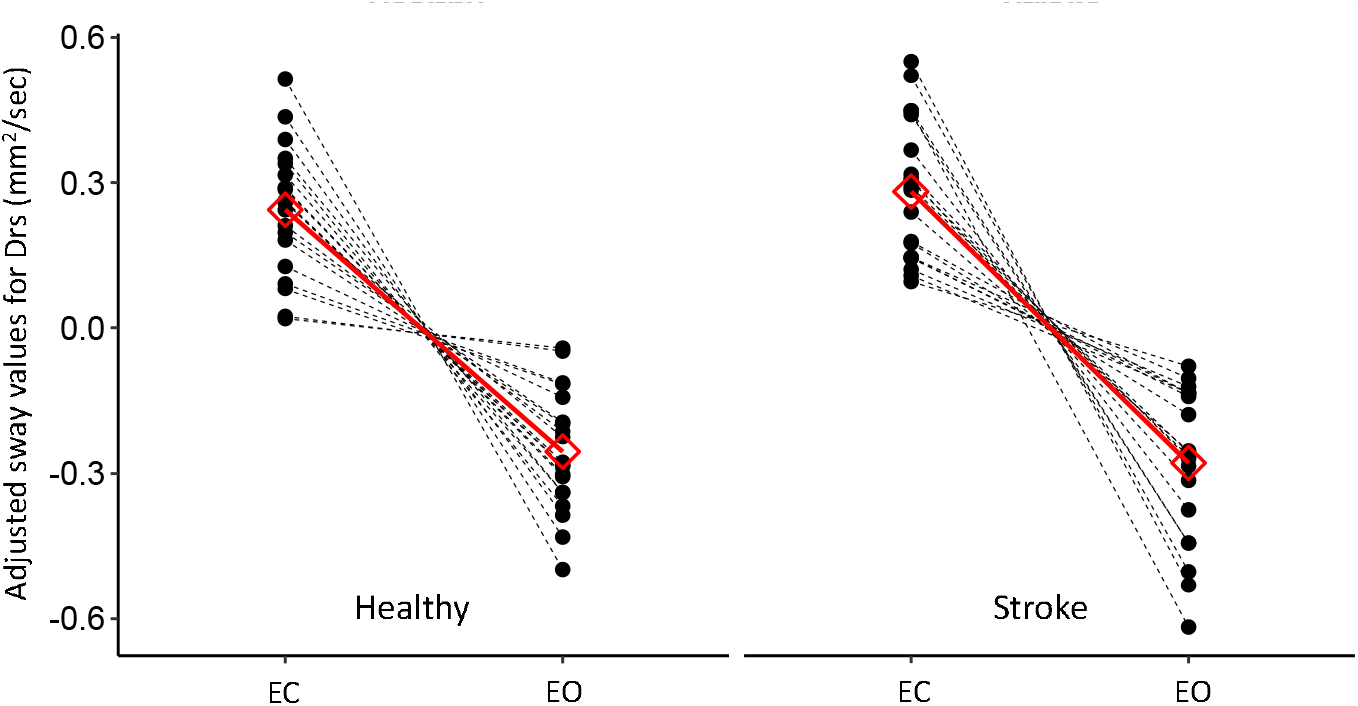
Between-group comparison of the participants’ individual (black) and the mean (red) responses to the visual condition. Results are presented as a within-subject effect (after adjusting for the random intercept) for the parameter Drs. EC is eyes closed, and EO is eyes open.

**Figure 2.**
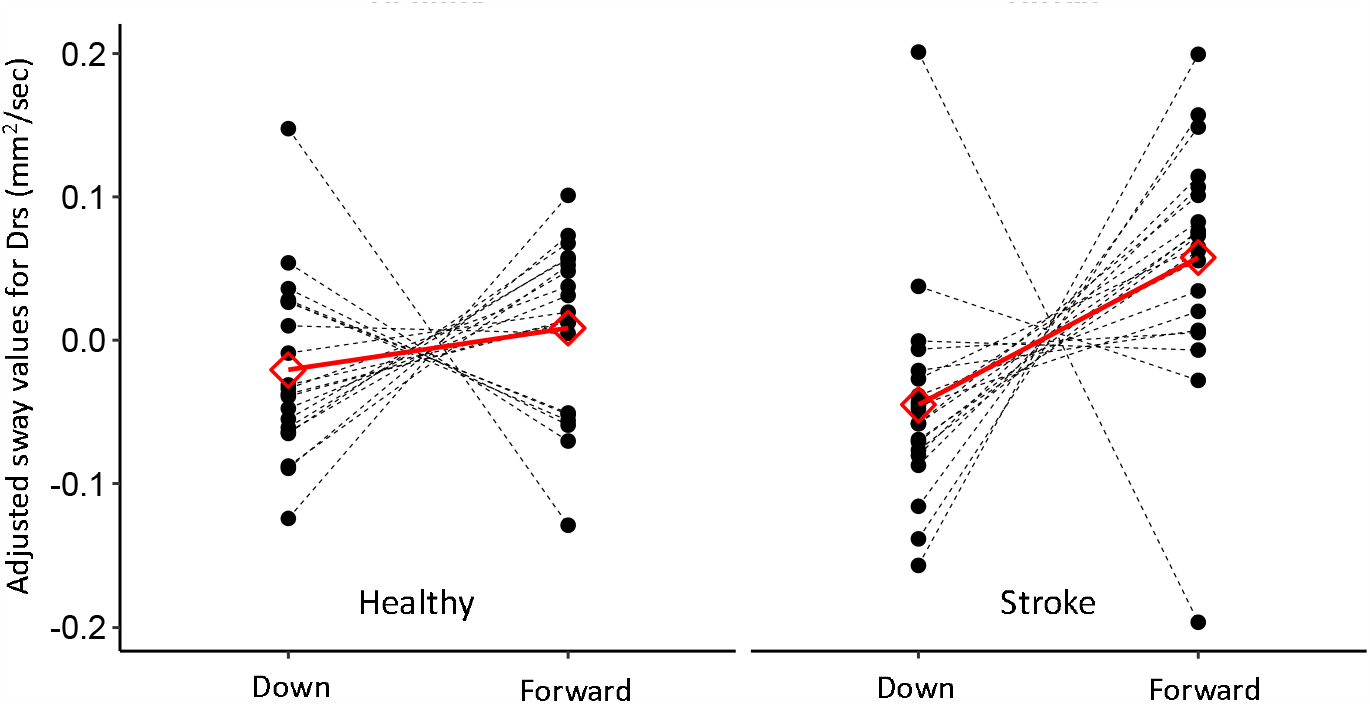
Between-group comparison of the participants’ individual (black) and the mean (red) responses to the gaze angle position. Results are presented as a within-subject effect (after adjusting for the random intercept), for the parameter Drs. While the response in the stroke group was greater (in terms of magnitude and the number of participants responding in the direction of the mean response) than that observed in the control (healthy) group, this difference was not significant for the parameter Drs; however, it was significant in other models.

Briefly, for all outcome measures we found the ‘Group’ and ‘Vision’ terms to be significant, revealing that mean sway values in the stroke group and with eyes closed, were significantly greater than the mean values in the healthy group and with eyes open, respectively.

The ‘Angle’ term was found to be significant in four out of the seven statistical models (four parameters), revealing that mean sway values in the FG condition were significantly greater than the mean values in the DWG condition. Nevertheless, a significant ‘Group’ by ‘Angle’ interaction (5/7 models) revealed that FG was indeed associated with increased sway values in all models, but that this effect was either observed only in the stroke group or was more pronounced in the stroke group. None of the other interactions were found to be significant. Importantly, none of the statistical models revealed a significant ‘Vision’-by-’Angle’ interaction, as would be predicted from our hypothesis.

Given that these results are clearly inconsistent with our hypothesis, we wanted to test whether the different gaze positions had any biomechanical effect, which is another possible explanation for the effect of DWG [14]. To do so we tested how the different gaze positions affected the relative weight born by the forefoot, i.e., the difference in percentage between the weight on forefoot and the weight on the backfoot. We concentrated on the AP direction because the down-tilt of the head was expected to shift the COM forward and down. The results of this model revealed a significant effect of only the ‘Angle’ term. Specifically, in the FG condition, the difference between the weight on forefoot and the weight on backfoot was 7.5% [1.8-13.2], and in the DWG condition, this difference was 11.2% [5.4-17]. This increase in weight born by the forefoot was statistically significant (p=8.8×10^−9^), indicating that DWG shifted the COM forward.

### Secondary results

Our secondary objectives were to investigate what gaze position (head angle) participants would choose to achieve the task’s goal (to stand as still as possible) and whether they would change this choice after being exposed to the experimental conditions.

First, we tested whether head angle was different between the first and second free-gaze trials. The results of this model revealed a 2.7° [-0.11-5.6] increase (upward) from the first to the second trial that was not significant (p=0.06). Next, we tested whether the head angle in the free-gaze condition was different from that angle in the FG and DWG conditions (EO only). The results of this model revealed that the head angle in the free-gaze condition was significantly different (p<0.001) than the head angle in the FG (−3.7° [1.7-5.7]) and the DWG (16.7° [13.2-20.1]) conditions (see Figure 3).

**Figure 3.**
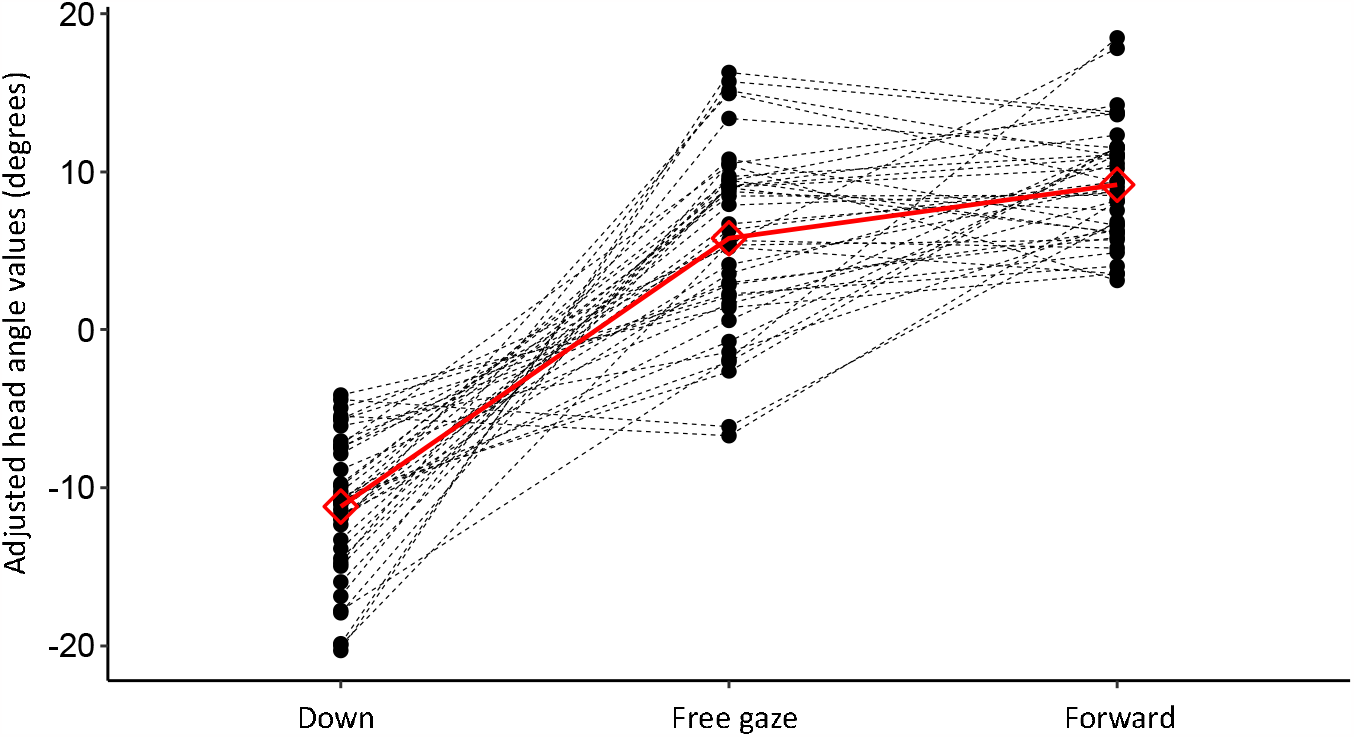
Between-angle comparison (eyes open only) of the participants’ individual (black) and the mean (red) head angles. Results are presented as a within-subject effect (after adjusting for the random intercept). All post-hoc pairwise comparisons were found to be statistically significant.

## Discussion

The main objective of this study was to provide evidence to support the hypothesis that the greater postural steadiness observed when a person gazes down is primarily a result of altered optic flow. The results of this study revealed that three factors had a negative influence on the ability of participants to attenuate their body sway (i.e., to control posture): lack of visual input, a previous stroke, and a straight (horizontal) gaze angle. Importantly, a significant ‘Vision’-by-’Angle’ interaction, which was required to support our main hypothesis, was not observed. The observed main effects of visual input and of stroke are consistent with current literature, including previous reports from our own laboratory [20,21] and will not be further discussed.

For postural control, the CNS uses sensory feedback from the somatosensory, vestibular, and visual systems [2]. Specific signals are not hardwired; instead, these signals are thought to be interpreted in a unified reference frame, allowing the CNS much needed flexibility. While this flexibility of the CNS allows adequate control with different gaze positions/angles, this does not mean that the magnitude of individual signals or the sensitivity of the sensory organs is unaffected by different gaze positions.

The “usual suspect” for such changes in magnitude is the retinal input, as it is sensitive to the visual structure of the environment (e.g., [31]). This is why several authors, who investigated the effect of different head and eye positions on postural control, tried to control, in one way or another, the retinal input in their investigations (e.g.,[14,15,18,31]). This approach simplifies the investigation but does not allow the researcher to determine whether this input has any beneficial or detrimental influence on postural control.

In this study, retinal input was matched between the two gaze angles only in the EC condition. Specifically, in the EC condition, retinal input was absent in both gaze angles. Thus, any effect of the gaze angle in this condition could not be attributed to the retinal input. On the other hand, any effect, or lack thereof, observed in the EC condition cannot serve to indicate what role, if any, is played by the visual input. Comparisons with previous reports might be confounded by many factors, including measuring equipment, stance width, instructions (i.e., the task), participants, etc. Naturally, any effect observed in the EO condition cannot serve this purpose because 1) it can originate from any sensory system/s, and 2) we cannot determine whether the visual input was similar between the two gaze angles.

To investigate whether visual input plays a role in the effect of DWG on postural control, we tested whether this input modulates the effect of DWG— namely, whether it changes this effect or not, as indicated by the interaction term in the statistical models. While several other authors [14,16,17,32] have used a similar approach, none directly tested the interaction term. Instead, they reported, for each visual condition, whether sway values were significantly different between gaze angles. While this approach is often acceptable, it does not necessarily indicate whether the magnitude of the effect changes [33]. The main results of this investigation do not point to any such modulatory effect of the visual input, indicating that visual input played no role in the effect of DWG on postural control.

With that being said, the fact that we found no modulatory effect of the retinal input should not be mistakenly construed as an indication that retinal input does not play a role in the effect of DWG on postural sway altogether, but instead serves as an indication that DWG can enhance postural control even in the absence of retinal input.

So, why did DWG enhance postural steadiness in the current investigation? While this experiment was not designed to cover all possible origins of the effect, there are several possible explanations. These may be used in the future to formulate new hypotheses and to design experiments to test them. Below, we briefly discuss two possible explanations:

1. One or more of the sensory organs that are used to detect body sway, or to calibrate the body scheme, become more sensitive in the downward gaze position.
2. The downward head position leads to greater biomechanical stability.

Starting with the latter possibility, neck flexion is expected to shift the COM downward and forward. In the DWG condition, we observed a 20° decrease in the head’s angle, indicating that participants flexed their necks although they were not specifically instructed to do so. Consistent with the above expectation, we observed an increase in the percentage of body weight born by the forefoot. While the lowering of the COM does increase mechanical stability (it reduces the moment around the ankle joint), the forward shift increases the distance (in the AP direction) of the projection of the COM from the center of rotation of the ankle joint and shifts it closer to the limit of the BOS. Both the greater distance from the ankle joint and the closeness to the limits of the BOS are thought to decrease stability [34]. In contrast to our observation, Buckley et al. [14] observed a backward shift of the COM with neck flexion, which is expected to have the opposite effect. Nevertheless, they observed an increase in body sway. If the effect of neck flexion were purely biomechanical, we should have observed an increase in body sway and Buckley should have observed a decrease. Thus, both our own and Buckley’s observations do not support a purely biomechanical effect.

The reason we observed a forward shift and Buckley a backward shift of the COM is likely a combination of biomechanical and physiological factors. Specifically, we measured 20° of neck flexion while Buckley used a 45° angle. The resulting forward displacement of the COM could be compensated for by increasing muscle activity in plantar flexors (i.e., ankle strategy), or by a compensatory backward shift of the COM using hip flexion (i.e., hip strategy). Greater neck flexion might pose additional/different constraints that could be better handled using a hip strategy [35]— that is, hip flexion with a backward shift of the pelvis, resulting in a backward shift of the COP. In fact, in a previous report from our laboratory [21], we observed that extreme neck flexion decreased postural steadiness, an observation consistent with Buckley’s report. The ankle-vs.-hip strategy (actually, Horak and Nashner proposed that these two reside on the same continuum) may explain the difference in the direction of COP shift, but not the different effects on body sway. Further, this biomechanical explanation accounts for only the effect observed for AP sway, not the effect observed for ML sway, both in the current and in previous reports [20,21]. Thus, a different or a complementary explanation is required.

Given that visual input and biomechanics cannot provide a complete explanation of the observed effect, it would be reasonable to assume that the vestibular and/or the somatosensory systems are involved. In fact, the vestibular system was suggested to play a central role in the negative effect of neck extension on postural control [36], which is a consistent finding in the literature [14,15,31,37]. Nevertheless, the effect of neck extension was suggested to result from positioning the otolith organ/s at the limit of their measuring range and shown to occur at extreme angles (40-50°). This is not the case here, and in any case, DWG did not disrupt but enhanced postural control. While the current experiment cannot support or refute the involvement of the vestibular organ in the observed effect, we believe that there are more likely explanations related to the somatosensory system.

The somatosensory system is comprised of many receptors throughout the body that could easily be affected by the downward head and/or eye position due to the stretching or relaxing of muscles, ligaments, skin, and joint capsules. Such changes can affect the sensitivity of these receptors, enhancing or reducing the ability of the CNS to formulate an accurate body scheme or to detect body sway. In fact, previous reports indicate that many factors, including the external environment, current and past position and motion, effort and many more, affect the perceived body scheme, its position, and motion (see several examples in [38]). One special case that comes to mind is the receptors in the muscles controlling the eyes. Both upward and downward eye positions and greater convergence angles were shown to enhance postural control [11,18], suggesting that in their stretched or contracted states, these muscles provide more accurate information that is used to attenuate body sway. While the eye position was not monitored in this study (due to obvious technical challenges), participants were specifically instructed to gaze onto the targets and in the eyes closed condition to imagine they were constantly looking at the designated target. This instruction was specifically given to match the eye positions in the EO and EC conditions.

In addition to the main effects observed, we also found a significant ‘Angle-by-’Group’ interaction in most models. This finding suggests that DWG enhanced postural steadiness to a greater extent in PwS than in healthy adults. This may indicate that PwS are more sensitive to this effect, which is consistent with a previous report [17]. By contrast, in a previous report from our laboratory [21] such an effect was not observed (except for the special case of extreme DWG, i.e., when participants gazed down to their own toes), but those participants were tested in a narrow-base stance. This fact may indicate that the magnitude of the effect is more related to the postural challenge than to a specific condition or population (see for example [30]). This possibility is consistent with a previous report [16] showing that DWG enhanced postural steadiness of healthy adults standing on one leg, an effect that disappeared when participants were tested in a two-legged stance.

DWG is a common clinical observation in unstable walkers. Previous investigations [39-46] indicate that when required to plan and guide subsequent steps, humans will gaze down, presumably to identify imminent or future foothold targets or obstacles. These observations, along with findings of greater tendency to gaze down in unstable walkers [17,43,47-49], might lead to the conclusion that unstable walkers are consciously trying to control stepping. By contrast, we presented evidence showing that DWG enhances postural control of both standing and walking adults [20,21] and that instability leads to DWG [50]. Whether used to plan and guide stepping, to control posture, or both, the underlying axiom is that DWG is used to gather visual information important for the task. Our current results suggest that this is not necessarily the case, and further emphasizes the notion that, by itself, spatiotemporal investigations of gaze behavior are insufficient to determine what visual information is being gathered, what purpose this information serves [51], or even whether a certain gaze behavior is aimed at gathering visual information at all. Nevertheless, DWG behavior in unstable walkers may allude to a compensatory strategy, a possibility that should be considered before targeting it in clinical settings.

## Data Availability

All data produced in the present study are available upon reasonable request to the authors

